# The Autonomous Cognitive Examination: Machine-Learning Based Cognitive Examination

**DOI:** 10.1101/2024.09.05.24313114

**Authors:** Calvin W. Howard, Amy Johnson, Sheena Barotono, Katharina Faust, Joseph Peedicail, Marcus Ng

## Abstract

**INTRODUCTION:** The rising prevalence of dementia necessitates a scalable solution to cognitive screening and diagnosis. Digital cognitive assessments offer a solution but lack the extensive validation of older paper-based tests. Creating a digital cognitive assessment which recreates a paper-based assessment could have the strengths of both tests.

**METHODS:** We developed the Autonomous Cognitive Examination (ACoE), a fully remote and automated digital cognitive assessment which recreates the assessments of paper-based tests. We assessed its ability to reproduce entire cognitive screens in a comparison cohort (n = 35), and the ability to reproduce overall diagnoses with an additional validation cohort (n = 11).

**RESULTS:** The ACoE reproduced overall cognitive assessments with excellent agreement (intraclass correlation coefficient = 0.89) and reproduced overall diagnoses with excellent fidelity (area under the curve = 0.96).

**DISCUSSION:** The ACoE may reliably reproduce the evaluations of the ACE-3, which may help in accessible evaluation of patient cognition. Assessment in larger population of patients with specific diseases will be necessary to determine usefulness.

## Background

Current projections estimate 150 million patients living with dementia worldwide by 2050, with 57 million as of 2019.^1^ The aging population presents a considerable diagnostic challenge as there are not enough physicians to examine every patient. The resulting healthcare strain is evidenced by the majority of dementia patients being undiagnosed,^2–5^ with those that do get a diagnosis taking 3-years or longer from symptom onset.^2,3,6–8^

Digital cognitive assessments (DCAs) offer a potential method to help improve diagnostic timelines.^9–11^ Among other things, two factors that influence the utility of DCAs at a population level are accessibility and generalizability.^11,12^

Some DCAs may sacrifice a degree of accessibility to better control testing conditions, increasing accuracy.^13,14^ Many DCAs achieve their exceptional performance by requiring specific hardware,^15–21^ in-office expert administrators,^13,15,16,18–20,22–24^ or by limiting variability by using mouse-and-keyboard questionnaires.^15,25–28^ Eliminating these requirements could improve accessibility, for non-affluent, rural, or cognitively impaired patients.

Other DCAs may focus on specific aspects of cognition which can best diagnose particular diseases.^9,15,18,22,23,28–33^ This approach yields high accuracy^9^ but may limit generalization to multiple domains of cognition or diseases. Although generalizing DCAs may reduce the ability to specialize on a particular disease, it may improve the ability to detect a range of diseases presenting in different cognitive domains.

Here, we validate the Autonomous Cognitive Examination (ACoE).^34,35^ It is hardware agnostic, self-administered, and naturalistically answered DCA. It allows patients to answer with speech, writing, and drawing, aiming to lower the cognitive function needed to accurately take the exam. Using 19 primary cognitive tasks, the ACoE estimates function in the main cognitive domains. The goal of the ACoE is not to classify patients and replace clinicians—it attempts to provide an estimate of their paper-based scores to support clinical decision making while enabling use of dementia detection algorithms.

We first compare the ACoE’s ability to estimate cognitive scores from Addenbrooke’s Cognitive Examination-3 (ACE-3) and subsequently validate its diagnostic discrimination against the Montreal Cognitive Assessment (MoCA).

## Methods

### Ethics Statement

The study has been conducted in accordance with the ethical standards. This study was conducted in accordance with ethical standards as described in the 1964 Declaration of Helsinki and its subsequent amendments. Approval was achieved by the Research Ethics Board of the Health Sciences Center, the University of Manitoba (#HS25666).

### Study Design

A two-period double crossover randomized-controlled trial was utilized to evaluate if the ACoE could reliably estimate ACE-3 score (Figure 1).^36^ Patients were randomized to receive either the ACoE or ACE-3 first, then returned 1-6 weeks later and received the other test. Subjects were randomized equally to ACoE or ACE-3 arms.

**Figure 1.**
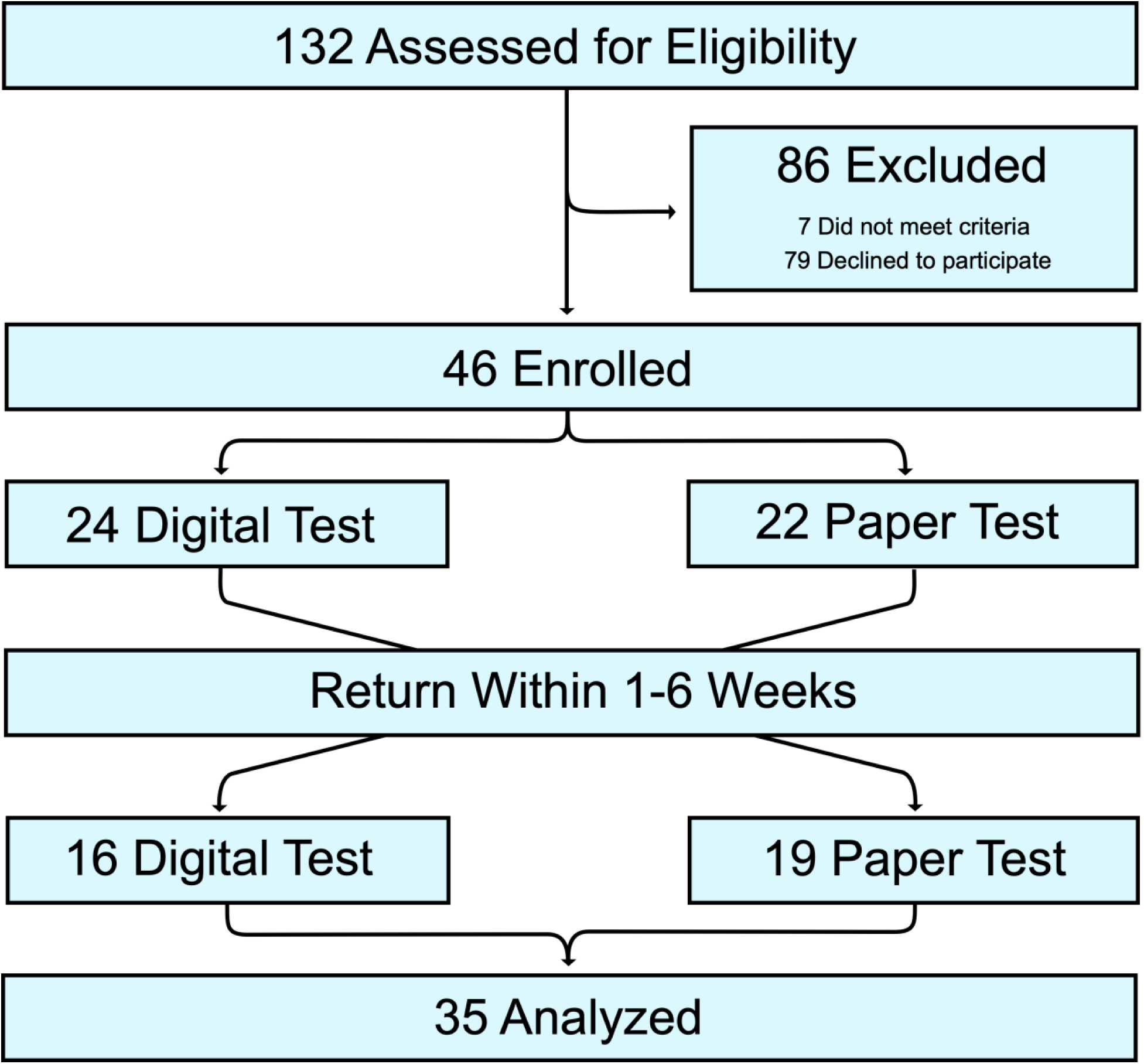
CONSORT Flow Diagram of Crossover Trial. 132 patients were evaluated for enrollment. 86 patients were excluded. 46 patients were enrolled in the trial, with 24 randomized to receive the ACoE first and 22 randomized to receive the ACE-3 first. Subsequently, each arm then crossed over and received the other test. 11 total patients were lost to follow-up. 35 patients completed the study and were analyzed.

This equivalently distributed learning bias and other confounds across each arm. Inter-test duration was limited to 1-6 weeks to control for time-, medication-, or pathology-related cognitive changes to balance for learning bias while also minimizing in disease or mental state between tests.^37^ No changes were made to the trial design. Only patients receiving the ACE-3 were randomized.

### Test Administration

Patients were tested in a quiet environment by a physician trained in cognitive examination. The ACoE was administered by touchscreen and microphone-equipped tablet. The ACoE administered itself to the patient without interference or prompting from the examiner. Caregivers were allowed to join but could not participate in the examination. ACoE responses were automatically scored and summated. The ACE-3 examination was performed via standard paper-based methods and evaluated using the ACE-3 scoring guideline.^37^ A trained physician examiner directly administered the examination, evaluated responses, and summated scores. Both tests consisted of 19 similar questions with 76 total sub-questions testing the same cognitive domains in the same order.

### Subjects – ACE-3

Our study cohort comprised patients from neurology clinics across the Health Sciences Centre, University of Manitoba. Patients indicating interest in clinical research were contacted by study team members via phone. Interested patients were screened for inclusion and exclusion criteria and enrolled. Upon enrollment, all patients underwent randomization to either arm of the study using a random number generator in a 1:1 ratio. This study was not blinded. At the first clinic visit, patients were again screened for inclusion/exclusion criteria by a physician. Patients or their caregivers provided written consent at the first clinic visit.

Patients with and without cognitive complaints were recruited. Inclusion criterion was English fluency. Exclusion criteria were acute medical conditions contributing to cognitive state, acute psychiatric disorders contributing to cognitive state, delirious states, or disabilities restricting ability to utilize screens, disabilities restricting ability to receive visual and auditory instructions, or developmental delay. Patients were recruited until powered sample size was achieved, with additional demographic variables available (Table 1).

**Table 1.** Patient Characteristics. Characteristics are split by randomization group.

| Characteristic | Group 1 | Group 2 |
| --- | --- | --- |
| Average age (years), median (IQR) | 46.3 (46) | 44.2 (39) |
| Age categories (years), N (%) |  |  |
| Under 25 | 0 (0) | 0 (0) |
| 25-45 | 9 (47.5) | 10 (62) |
| 45-65 | 9 (47.5) | 3 (18) |
| Over 65 | 1 (5) | 3 (18) |
| Sex, N (%) |  |  |
| Male | 8 (42) | 9 (56) |
| Female | 11 (58) | 7 (44) |
| Ethnicity, N (%) |  |  |
| White | 12 (64) | 7 (43) |
| Indigenous | 3 (16) | 2 (13) |
| Indian | 1 (5) | 3 (18) |
| Filipino | 1 (5) | 2 (13) |
| African | 1 (5) | 0 (0) |
| Eastern European | 1 (5) | 2 (13) |
| Education, N (%) |  |  |
| Less than Secondary | 0 (0) | 2 (12.5) |
| Secondary | 15 (79) | 12 (75) |
| Post-Secondary | 3 (16) | 2 (12.5) |
| Graduate or Professional | 1 (5) | 0 (0) |
| Employment Status, N (%) |  |  |
| Unemployed | 7 (37) | 4 (25) |
| Employed | 12 (63) | 12 (75) |
| Diagnosis, N (%) |  |  |
| Neurologically Healthy | 5 (26) | 6 (38) |
| Mild Cognitive Impairment | 4 (21) | 3 (18) |
| Probable Alzheimer Disease | 2 (11) | 1 (6) |
| Epilepsy | 8 (42) | 6 (38) |

### Subjects – MoCA

Patients were enrolled from neurology clinics across the Health Sciences Centre, University of Manitoba. Patients indicating interest in clinical research were contacted by study team members via phone. Interested patients were screened for inclusion and exclusion criteria and enrolled. These patients were not randomized and were not included in evaluation of scoring reliability just in evaluation of diagnostic reliability.

Patients or their caregivers provided written consent at the first clinic visit. 11 (5 neurologically healthy, 2 mild cognitive impairment, 4 Alzheimer Disease) patients received the MoCA. Average age was 61 ± 16 years, ranging from 33 to 82 years of age.

### The ACoE

The ACoE receives user input via any hardware device with an internet connection, microphone and touchscreen. The ACoE questions are answered via microphone and touchscreen, although a keyboard and mouse can be used. The user proceeds through an examination from end-end which automatically administers voiced instructions with closed captioning (Figure 2).

**Figure 2.**
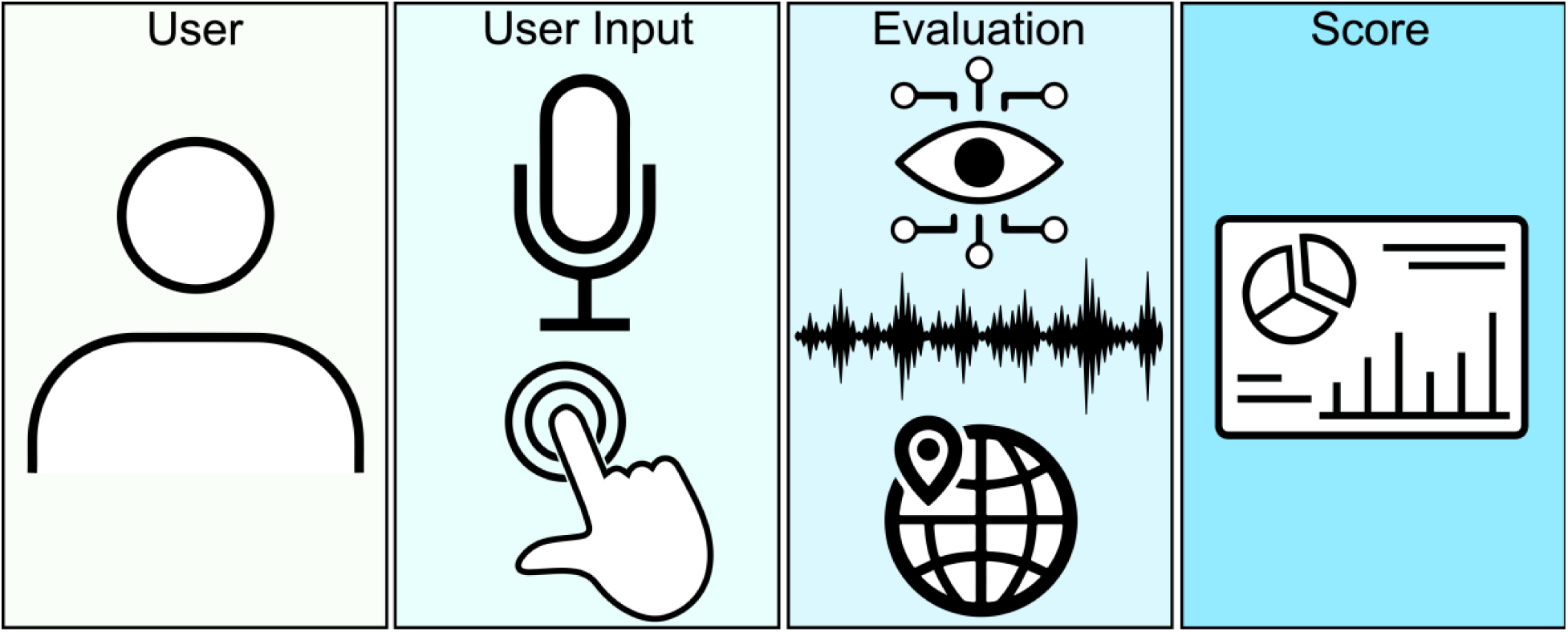
The workflow of the ACoE. The examination begins with an individual user, either alone or accompanied by a caregiver. User inputs are primarily via microphone and touchscreen, although mouse and keyboard are allowed. User inputs are evaluated using a host of 79 algorithms. These algorithms span three primary domains: computer vision for evaluation of drawings, natural language processing for evaluation of speech, and expert algorithms to evaluate all other inputs, such as geolocating a patient to verify orientation to space and time. Scores output by the individual evaluation algorithms are summated, providing the final total score as well as the scores for cognitive domains of memory, language, fluency, visuospatial, and attention.

The ACoE consists of 19 questions within the primary domains of memory, language, fluency, visuospatial, and attention function. Attention comprises executive function and actual attention. This classification system of the cognitive functions is modeled after the ACE-3.^37^ A table of each question and associated primary cognitive domain are available (Supplementary Table 1).

For each question, the patient is given unrestricted time to answer. The patient receives specific instructions both visually and by audio at the start of each question. The instructions were allowed to be repeated up to 3 times, but no further assistance is provided. 13 questions are answered with speech, 3 with touchscreen inputs of drawing or manipulating on-screen objects, 2 with dropdown menus, and 1 with typing.

At the end of the examination, total score and cognitive domain scores are calculated. There are 5 memory questions totaling 26 points, 1 fluency question totaling 14 points, 7 language questions totaling 40 points, 3 attention questions totaling 18 points, and 3 visuospatial questions totaling 16 points. The total score totals 100 points.

### ACoE Scoring

A unique algorithm was developed for each of the 19 primary questions, including sub-questions. This resulted in 76 unique algorithms, which have been previously described (Figure 2).^34,35,38,39^ In brief, each algorithm corresponds to how one question on the ACE-3 is scored and attempts to estimate the scoring of the ACE-3 for the corresponding question. These span three primary algorithmic domains: computer vision, natural language processing, and expert algorithms.

The 3 computer vision algorithms enable testing of visuospatial drawing tasks. For example, the overlapping infinity copy, cube copy, and clock drawing test. These rely upon a custom convolutional neural network created for the ACoE, the SketchNet.^39^ The questions these are responsible for represent 8/100 ACoE points.

The 48 natural language processing evaluates spoken answers, assessment of speech quality, sentence structure, and word pronunciation. These allow evaluation of spoken memory, language, fluency, executive, and visuospatial tasks. The tasks these are responsible for are immediate recall, mental arithmetic, delayed word recall, phonemic list generation, semantic list generation, semantic memory, sentence writing, word repetition, sentence repetition, naming, reading aloud, counting, identifying and partially obscured objects for simultanagnosia. The questions requiring natural language processing comprise 68/100 ACoE points.

The 25 expert algorithms allow evaluation of complex tasks which do not easily fit into machine learning approaches. For example, we developed a novel two-step algorithm to evaluate a patient’s orientation to their location in space. Natural language processing is used to understand the patient’s spoken answer. Then, geolocation is used to triangulate the patient’s true location, which is subsequently compared to their true location. If a patient is within an acceptable margin of error, after accounting for the curvature of the globe, they are evaluated as being oriented to their location in space. Similar algorithms are used to assess orientation to time, ability to follow on-screen commands, recognize objects, or recall with prompting. The questions requiring expert algorithms compose 24/100 ACoE points.

### ACoE Deployment

The ACoE is hosted on Amazon Web Services to provide accessibility to roughly 75% of the globe. The ACoE leverages a cloud-based format to enhance accessibility for patients, allow physicians to test patients regardless of their location, and to provide secure storage of data. Patients access the ACoE through links which are sent to them by an ACoE administrator. Each link is specific to the given patient and becomes inactive after use. Each link is validated upon use and invalid links are rejected access. The patient then receives the ACoE and results are sent via encryption to the scoring server which is stored on a private subnet. The scoring server then returns patient scores in an encrypted manner to the administrative user’s database. This allows the clinician using the ACoE’s administrative platform to view patient results as they are completed. The raw files for each patient are stored in an anonymized format on and encrypted and private database. The ACoE and administrator platform are Health Information Privacy Protection Act compliant. A diagrammatic representation of the process is provided (Figure 3).

**Figure 3.**
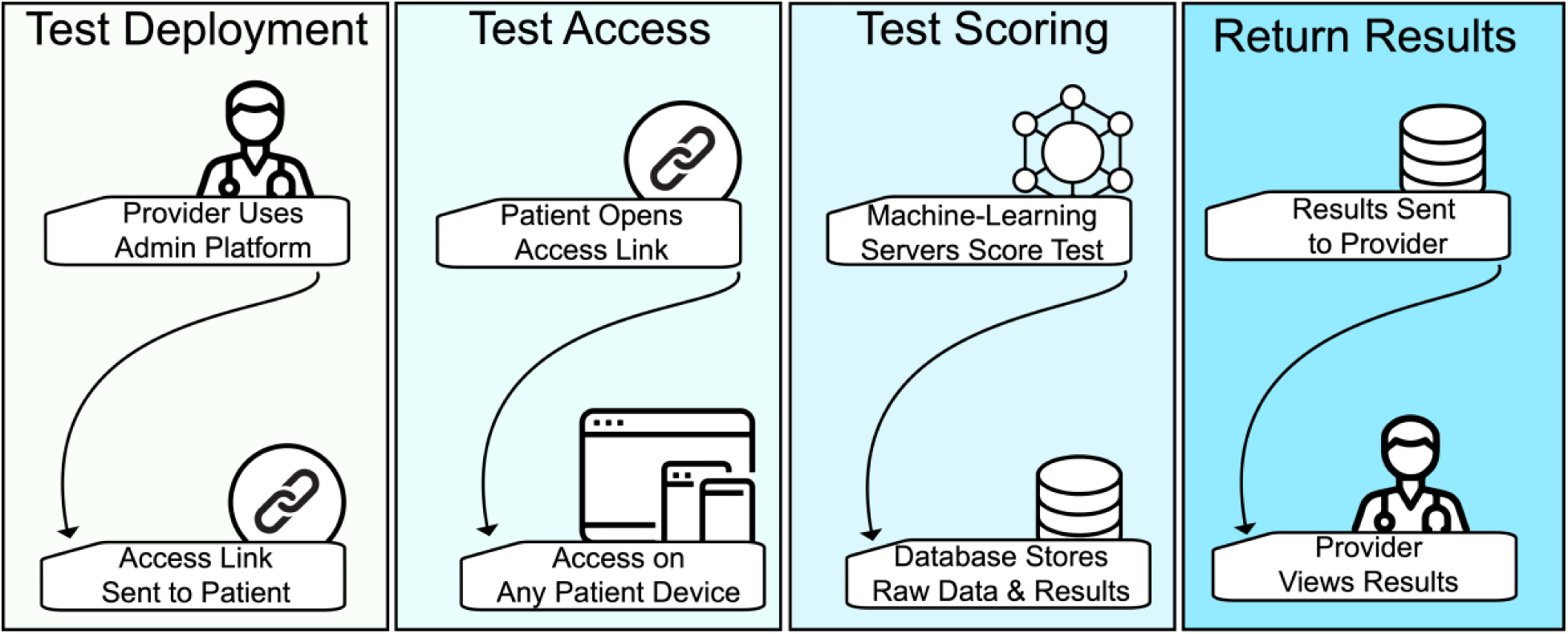
Utilization of the ACoE. Test Deployment) The testing process is initiated by the healthcare provider. The administrative platform is used to generate an access link for a patient, and is sent to the patient. Patients can also access ACoE links directly for self-. Test Access) Patients click the access link to access the exam, which runs on all devices. Test Scoring) Raw patient responses are sent to the scoring server. The scoring server receives the patient files, preprocesses them, and administers the evaluation algorithms to them. Summated patient scores are then stored in a database. Return Results) Summary results, broken down by cognitive domain and overall performance are sent to the administrative platform so the healthcare provider can view the patient’s results.

### Randomized Controlled Trial Sample Size

Power analysis for the randomized controlled trial was based upon established power analysis for ICC-based studies. Prior comprehensive power analyses of the ICC have been performed.^42^ Using these established power analyses, we require a sample of 35 patients to achieve 80% statistical power.

### Evaluation of Scoring Reliability

To evaluate reliability of the ACoE scores, the ICC was used to compare the similarity of scores between each patient’s ACoE and ACE-3 scores. To evaluate systematic deviances in group-level scores, we compared the central tendency of ACoE versus ACE-3 score with Wilcoxon signed-rank testing, a paired evaluator of the median.

### Diagnostic Reliability Power Analysis

Diagnostic reliability sample size was guided by receiver operating characteristic (ROC) area under the curve (AUC requirements). 16 positive and 16 negative cases were required to achieve 80% statistical power. This was based on worst-case-scenario requirements of an AUC of 0.70, while 3 positive and 3 negative cases were required for 80% statistical power for an AUC of 0.90, as per the Hansen and MacNeily formula.^43^

### Definition of Cognitive Impairment

Patients received a label of cognitive impairment based on established cutoffs for each test: 26/30 on the MoCA and 83/100 on the ACE-3.^37,44,45^

### Evaluation of Diagnostic Reliability

An ROC was constructed comparing the ACoE classifications to ACE-3 (n = 35) and MoCA (n = 11) classifications. AUC was calculated as a metric of reliability in diagnosis. Youden’s J was calculated to derive optimal classification threshold, enabling direct comparison of thresholds between ACoE and ACE-3.^46^ To assess the confidence with which a classification of impairment can be made, bootstrapped sensitivity and specificity were calculated at all ACoE scores. Specifically, patients were resampled with replacement 10,000 times, which is a reliable method of generated confidence intervals.^47,48^

### Covariates Influencing Discrepancy Between Scores

To identify patient factors which might be influencing the discrepancy between ACoE and ACE-3 scores, we performed a series of multivariate regressions relating the primary patient covariates of age, sex, educational status, and employment status on difference between ACE-3 and ACoE score.

### Covariates Influencing ACoE Score

We derived adjustment factors for the ACoE score dependent upon covariates. We related the primary patient covariates of age, sex, educational status, and employment status on ACoE scores. Given sample size, one large multivariate equation with all covariates was not used given it reduced the statistical power of each individual covariate. Thus, each covariate of interest was related to ACoE scores while controlling for cognitive status. This was performed with an only least squares multivariate regression.

### Adjustment for Patient Age

An adjustment factor which accounts for patient age will allow generalization of the ACoE score across age groups. We modelled the relationship of age upon ACoE scores after accounting for cognition (Supplementary Equation 1). The coefficient of age from this formula can be used to adjust ACoE scores for age (Supplementary Equation 2). The model relating cognitive status and age to ACoE scores was fit using the pooled cohort of MoCA and ACE-3 patients given a larger sample of older patients in the MoCA cohort.

### Statistics

All analyses were performed in Python. Central tendency, correlation, normality, and general linear model analyses were performed with Statsmodels. Power analyses were performed in accordance with established techniques, using a nomogram for intraclass correlation coefficient and a Python implementation.^43^ Intraclass correlation coefficient was calculated with the Pingouin package.^49^

Two methods of intraclass correlation were performed. The two-way random effects model is commonly employed to evaluate agreement between clinical evaluations and scales, and is what we use to derive our primary ICC results.^50^ We focus on evaluating the consistency of the two raters, the ACoE and the ACE-3. To perform sensitivity analyses, we use a much more conservative model,^50^ a one-way random effects model measuring the absolute agreement of the ACoE with the ACE-3. 95% confidence intervals were calculated for each ICC.

Subsequently, we use a one-way random effects model a two-way random effects, allowing evaluation of multiple raters and focusing on their consistency was used to evaluate the primary ICC results. Subsequently, we used a less Following this, reliability tests were performed with a subject (one-way random effects, absolute agreement, single-rater)

Two-way mixed effects, consistency, multiple raters/measurements

Spearman correlation was used for ordinal data. Paired Wilcoxon tests was employed for ordinal data and when normality was violated as measured by the Shapiro-Wilks test. Multiple comparisons were corrected with Bonferroni correction.

## Results

### Randomized Controlled Trial Recruitment Results

132 patients were assessed for eligibility. 79 patients denied enrollment and 7 did not meet inclusion criteria. 46 patients were randomized, with 24 patients randomized to receive the ACoE first (Group 1) and 22 to receive the ACE-3 first (Group 2). 11 patients were lost to follow-up. 35 patients completed the study and were analyzed. The patient flow chart is available (Figure 1). There were no adverse outcomes reported.

The 35 patients analyzed patients had a mean age of 45.3 ± 12.8 years (Male N=17, Female N=18). Cohort demographics comprised Indigenous Canadian (N=5), Indian Canadian (N=4), Filipino Canadian (N=3), African Canadian (N=1), Eastern European (N=3), and Caucasian (N=19) individuals. Further patient characteristics available (Table 1).

We next evaluated for appropriate randomization. Cognitive scores were not significantly different between the two arms (Wilcoxon test, p > 0.05, Supplementary Figure 1). Nor were there significant differences in the number of patients in each arm (Chi-squared, p > 0.05).

### Reliability of the Autonomous Cognitive Examination

We first evaluated the overall correlation of the ACoE’s assessment of cognition to the ACE-3’s assessment (Figure 4A). There was a significant positive correlation between the two tests (Spearman’s Rho = 0.87, p < 0.0001). To more conservatively assess how well each test administered scores on a patient-by-patient basis, we investigated the inter-rater reliability (Figure 4B). We found the ACoE and ACE-3 had significant inter-rater reliability (ICC = 0.89, 95% CI 0.79-0.95, p < 0.0001). We tested the reliability of this result using the most conversative ICC analysis, and found the agreement was still significant (p < 0.001).

**Figure 4.**
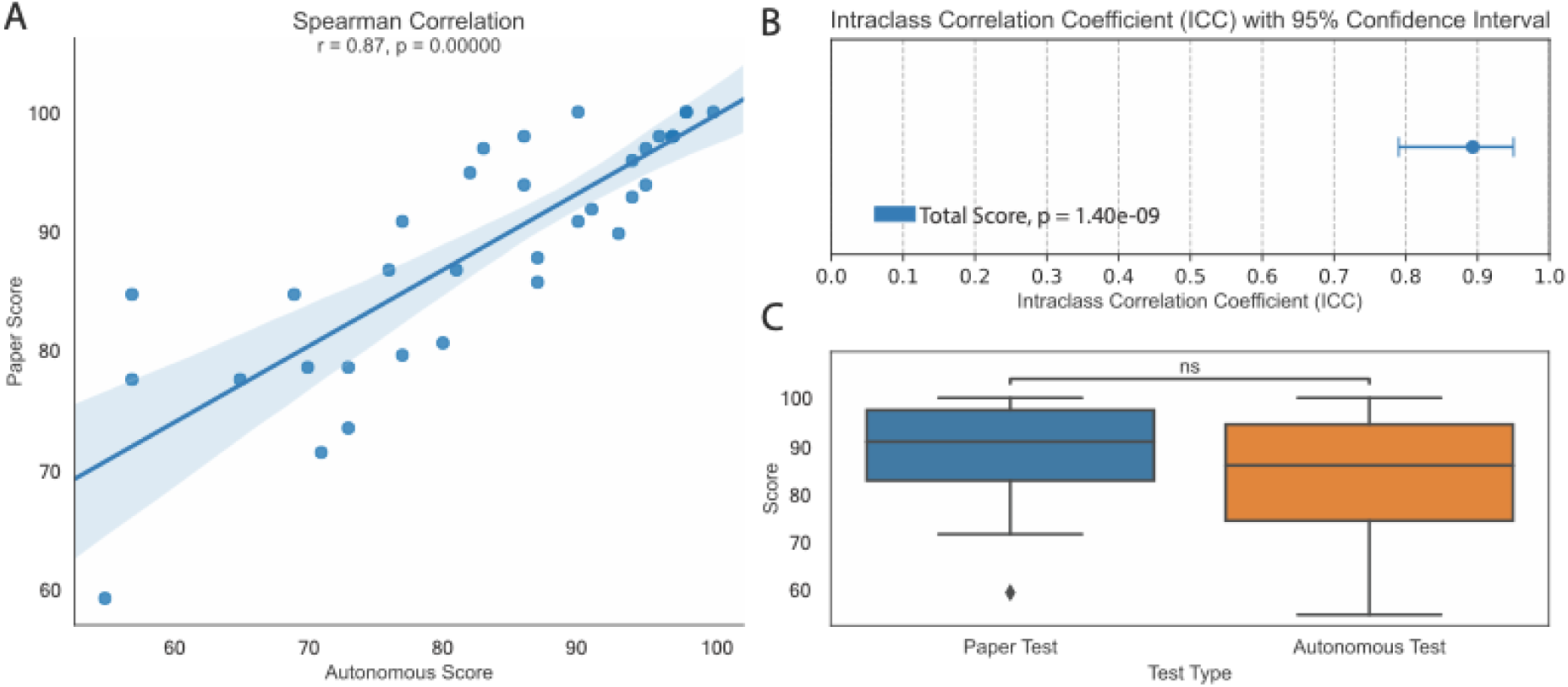
Reliability of Total Score Assessment. A) Spearman correlation of ACoE to ACE-3 demonstrates high correlation of patient scores between the tests (Rho = 0.87, p < 0.001). B) The ACoE reliably rates patients similarly to the ACE-3 (ICC = 0.91, p < 0.00001). 95% confidence interval of the ICC is presented. C) Wilcoxon test of ACoE relative to the ACE-3 demonstrates there is no significant difference between the median scores. Scores are presented as percent.

To assess for significant bias in overall scoring, we compared the median score of the ACoE and ACE-3 (Figure 4C). Distributions were found to be non-normal (Shapiro-Wilk, p < 0.05), and non-parametric testing found no significant difference between test scores (Wilcoxon Test, p > 0.05). Reliability tests also revealed no difference between the means when tested parametrically (T-test, p > 0.05).

### Reliability of Cognitive Domain Assessment

We next investigated the reliability of each cognitive domain’s assessment (Figure 5A). The ACoE was significantly reliable for all cognitive domains, including attention (ICC = 0.74, p < 0.00001), language (ICC = 0.89, p < 0.00001), memory (ICC = 0.91, p < 0.00001), fluency (ICC = 0.74, p < 0.00001), and visuospatial function (ICC = 0.78, p < 0.0001). To test the reliability of these results, we utilized the most conservative form of the ICC and assessed each domain again. All results remained significant, including attention (p < 0.0001), language (p < 0.0001), memory (p < 0.0001), fluency (p < 0.0001), and visuospatial function (p < 0.001).

**Figure 5.**
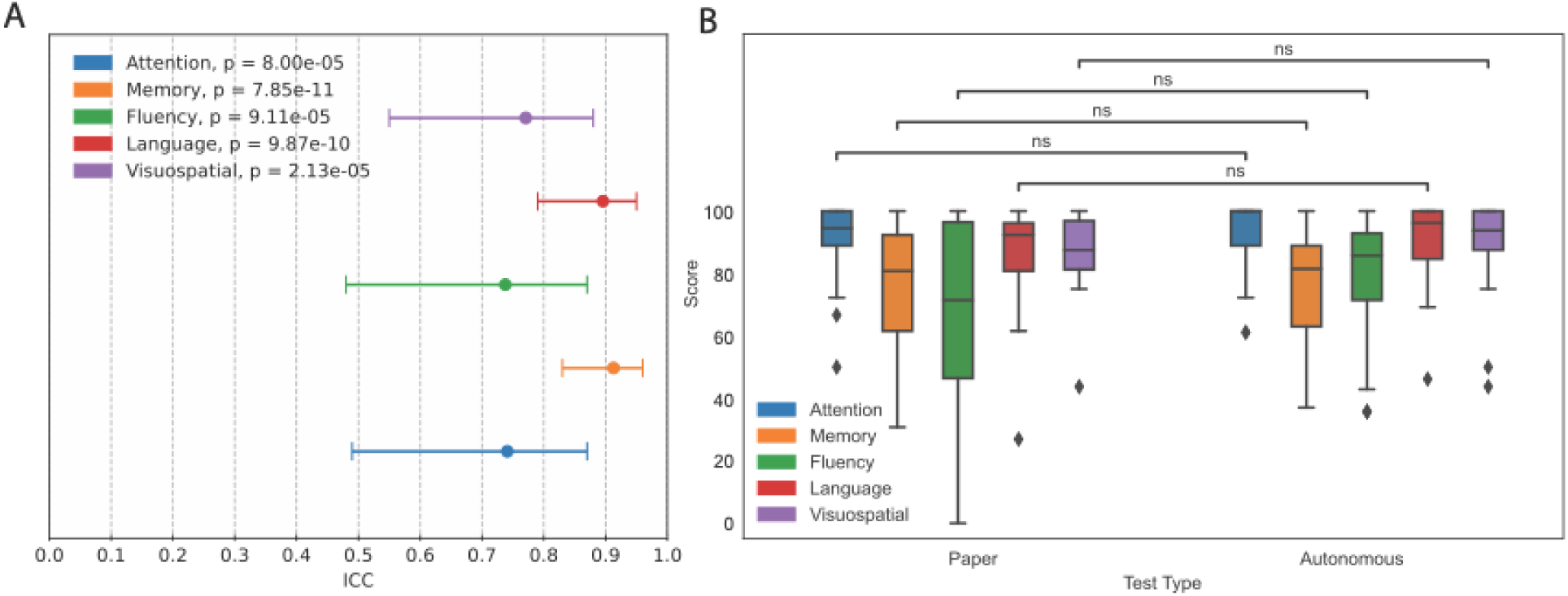
Reliability of Cognitive Domain Assessment. A) Reliability of ACoE cognitive domain evaluations ranges from high to very high as measured by ICC. 95% confidence intervals are presented with each ICC. B) Wilcoxon test demonstrates no significant difference between central tendencies of cognitive domains between either test. Scores are presented as percent.

We next compared the distribution of cognitive domain scores between the two tests (Figure 5B). Neither ACoE nor ACE-3 cognitive domain scores were normally distributed (Shapiro-Wilk, p < 0.05). Nonparametric testing of the medians revealed no significant differences between ACoE and ACE-3 in all domains, including attention (Wilcoxon Test, p > 0.05), memory (Wilcoxon Test, p > 0.05), visuospatial function (Wilcoxon Test, p > 0.05), fluency (Wilcoxon Test, p > 0.05), and language (Wilcoxon Test, p > 0.05). Sensitivity testing with parametric evaluation again revealed no differences in attention (T-test, p > 0.05), memory (T-test, p > 0.05), visuospatial function (T-test, p > 0.05), fluency (T-test, p > 0.05), and language (T-test, p > 0.05). Means, standard errors, and medians are available in the supplements (Supplementary Table 2).

### Reliability of Underlying Algorithms

As a metric of accessibility, we evaluated if the algorithms used to enable naturalistic completion of the exam were reliable (Figure 6A). We found the algorithms were all significantly reliable, including computer vision (ICC = 0.67, p < 0.001), natural language processing (ICC = 0.86, p < 0.0001), and the expert algorithms (ICC = 0.82, p < 0.0001). Results were stable after repetition with the more conservative ICC, including computer vision (p < 0.05), natural language processing (p < 0.0001), and the expert algorithms (p < 0.0001).

**Figure 6.**
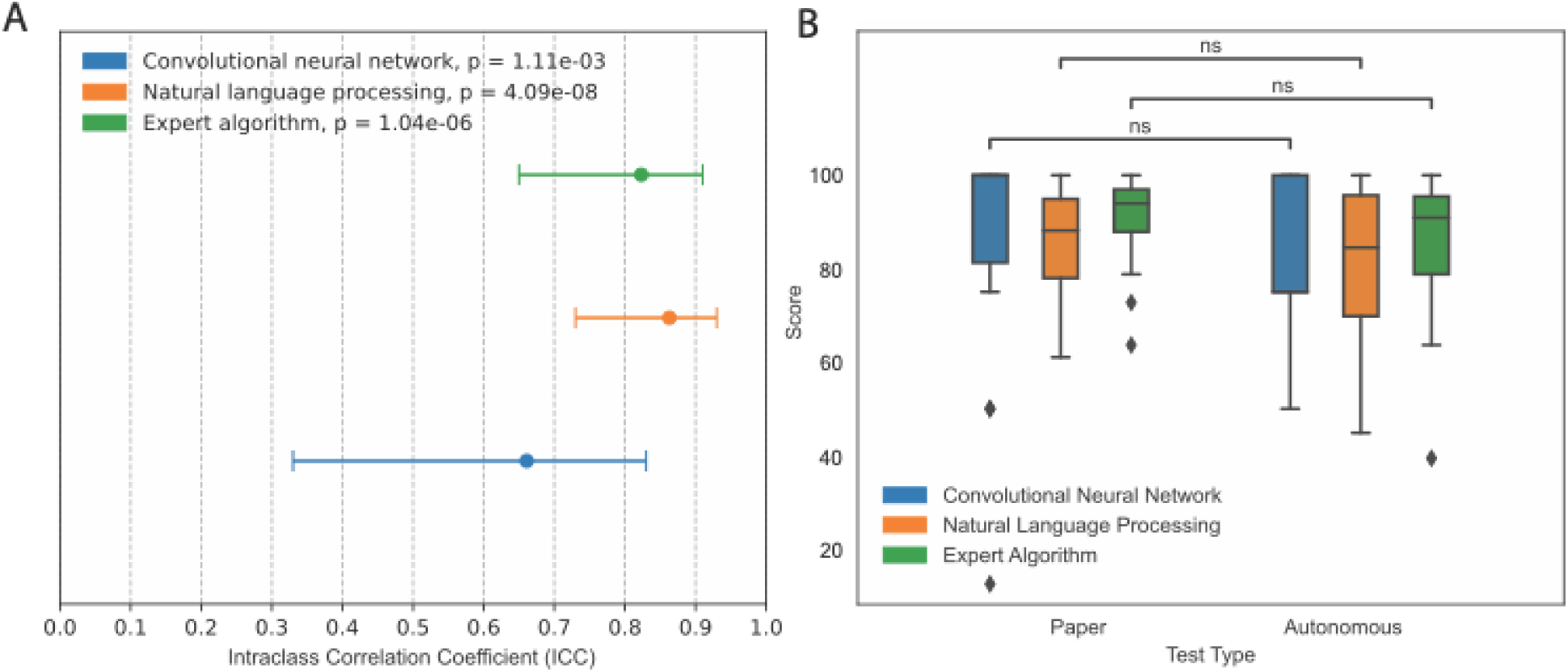
Reliability of the algorithms used to assess patient inputs. A) Reliability of the algorithms used in the ACoE range from high to very high as measured by ICC. 95% confidence intervals are presented with each ICC. B) Wilcoxon test demonstrates no significant difference between central tendencies of any test (p > 0.05). Scores are presented as percent.

We next compared the distribution of algorithm-based question scores between the two tests (Figure 6B). Neither ACoE nor ACE-3 scores were normally distributed (p < 0.05). Nonparametric testing revealed no significant difference across the algorithmic questions and their paper-based equivalents for computer vision (Wilcoxon test, p > 0.05), natural language processing (Wilcoxon test, p > 0.05), and the expert algorithms (Wilcoxon test, p > 0.05). Sensitivity testing with parametric evaluation also demonstrated no difference in natural language processing (T-test p > 0.05), computer vision (T-test p > 0.05), nor the expert algorithms (T-test p > 0.05) and their paper-based counterparts. Means, standard errors, and medians are available in the supplements (Supplementary Table 3).

We also evaluated each of the 19 questions for differences in algorithmic scoring versus their paper-based counterparts and found none (Supplementary Table 4).

### Identifying and Adjusting Covariates Influencing ACoE Scores

To assess accessibility, we evaluated if any patient demographic factors were associated with worse performance on the ACoE. Controlling for cognitive status, we performed a series of multivariate regressions relating each demographic variable to ACoE scores (Supplementary Figure 2). Only age was significantly negatively related to ACoE score (β_age_ = -0.22, p < 0.01). Interestingly, age did not interact with cognitive status to compound the effect of either upon ACoE score (β_interaction_ = -0.08, p > 0.05). This was specific to the ACoE, as age was not significantly related to ACE-3 score, nor was any other demographic variable (Supplementary Figure 3).

### Statistically Removing Age Bias

Given age was associated with worse performance, regardless of cognitive status, the ACoE demonstrated a slight bias against elderly individuals (Supplementary Figure 4A). We next evaluated if it is possible to remove this bias from patient scores post-hoc by accounting for the effect of age. After regressing cognitive status out of ACoE scores (Supplementary Figure 4B), age alone was significantly negatively correlated with ACoE scores (r = -0.37, p < 0.001). Then, we adjusted each patient’s score for their age and repeated the analysis (Supplementary Figure 4C). The adjusted scores removed the bias of age (r = 0.001, p > 0.05).

### ACoE Classifies Patients Similarly to Paper-Based Tests

Next, we assess the reliability of the age-adjusted ACoE. First, we replicate the evaluation of overall cognitive score reliability (Figure 7A). The reliability between the adjusted ACoE and ACE-3 was significant (ICC = 0.88, p < 0.0001). This was robust to the more conservative evaluation of ICC (p < 0.0001). The reliability within each cognitive domain remained significant, regardless of which ICC was used (p_max_ < 0.00001).

**Figure 7.**
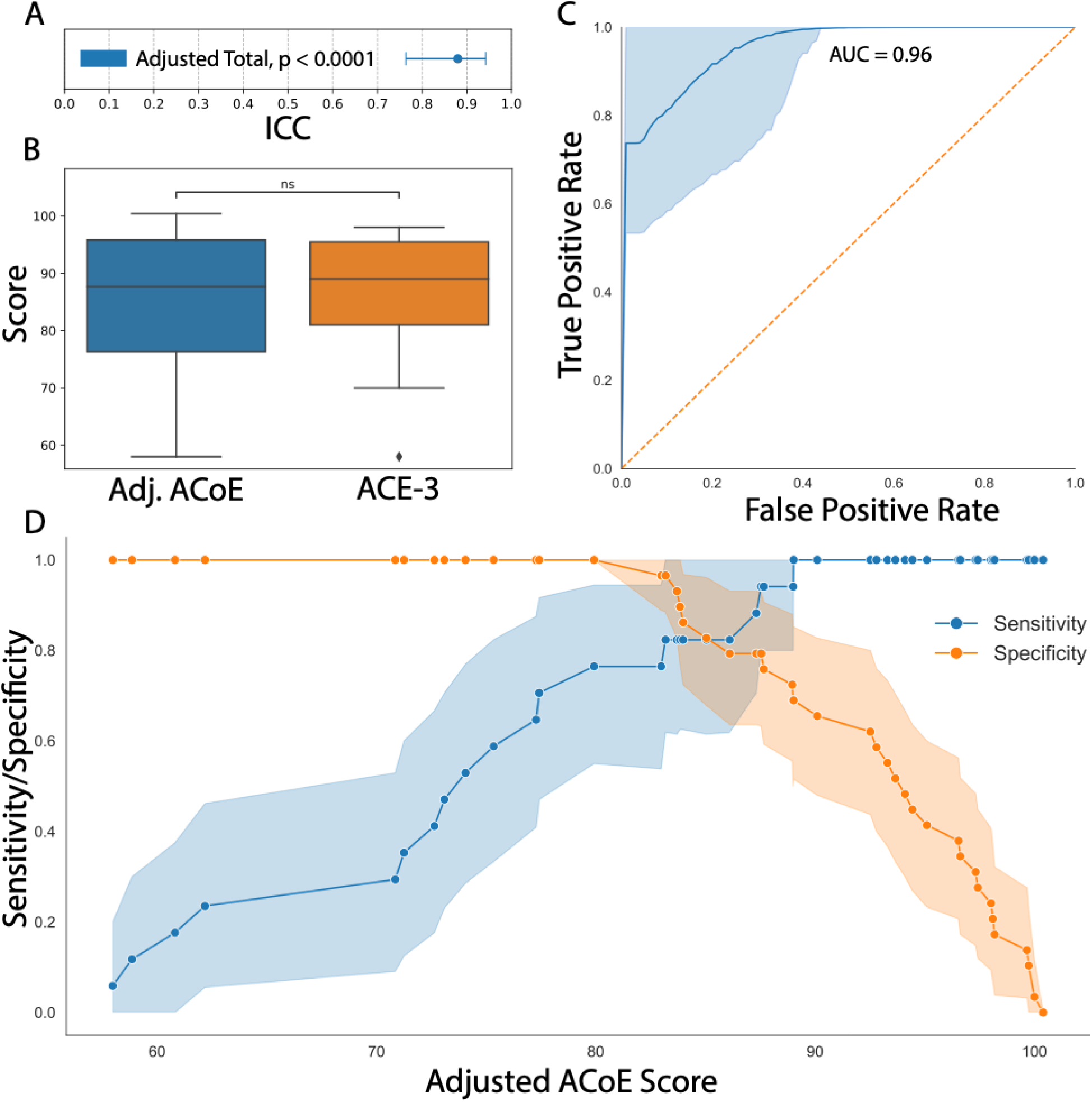
Age-adjusted ACoE Score Maintains Reliability and Diagnostic Accuracy of Paper-Based Tests. A) Reliability of age-adjusted ACoE Scores remains high. ICC of age-adjusted ACoE score to patients with ACE-3 score (n = 35) is 0.88 (95%CI 0.77-0.95). B) There is no systematic bias between age-adjusted ACoE scores and ACE-3 scores. Test of medians between age-adjusted ACoE score and ACE-3 revealed no difference (p > 0.05). C) ACoE maintains diagnostic consistency with paper-based tests (ACE-3 and MoCA). ROC AUC of the age-adjusted ACoE is 0.96 compared to MoCA and ACE-3. D) Score-specific sensitivity and specificity of the ACoE. Below of a score of 83%, the ACoE achieves a specificity of 1.0 (95%CI 1.0-1.0). Above a score of 89%, the ACoE achieves a sensitivity of 1.0 (95%CI 1.0-1.0).

We also repeated evaluation of the distributions of the two tests (Figure 7B). Nonparametric testing revealed no significant differences (Wilcoxon text, p > 0.05). Sensitivity evaluation with parametric tests also revealed no significant difference (T-test, p > 0.05). There were no significant differences between cognitive domain distributions, regardless of parametric or nonparametric testing (p_min_ > 0.05).

Finally, we compare the overall classification adjudicated by the ACoE, compared to the ACE-3 and an additional validation cohort of patients who took the MoCA (n = 11). First, an ROC was constructed to compare ACoE classification to ACE-3 and MoCA classifications, demonstrating an AUC of 0.96 (p < 0.0001). Youden’s J revealed the binary classification optimal threshold was 83%, the same diagnostic cutoff as the ACE-3.^37^ The AUCs were exceptional for both the ACE-3 alone (AUC = 0.98, p < 0.001) or the MoCA alone (AUC = 0.91, p < 0.001).

To better understand how ACoE classification accuracy varies across possible scores, we constructed sensitivity-specificity curves (Figure 7D). We find under an age-adjusted ACoE score of 83%, the test achieves a confident specificity of 1.0 (95% CI 1.0-1.0, p < 0.0001). Above an age-adjusted score of 89%, the test achieves a confident sensitivity of 1.0 (95% CI 1.0 – 1.0, p < 0.0001). This was repeated for positive and negative predictive values, which again confirmed 83% was the threshold for detecting impairment (Supplementary Figure 5). Control analysis using the unadjusted ACoE score revealed similarly significant diagnostic ability and sensitivity/specificity (Supplementary Figure 6).

## Discussion

### Validation of the ACoE

The primary concern in this was that in recreating an entire cognitive examination process from scratch, it may fail to accurately reproduce the nuance of in-person cognitive examinations.^37,51–56^ There are many sources of potential inaccuracies: poor instructions, poor transduction of responses, and poor evaluation of responses—each of these could occur within any of our 76 algorithms. However, in this small but to-power study, we find the ACoE can highly reliably reproduce the overall scores of the ACE-3,^57^ the evaluation of ACE-3 cognitive subdomains, and the overall classifications of the ACE-3 and even MoCA. Moreover, the ACoE has the exact diagnostic cutoff of the ACE-3.^37^ This suggests the ACoE may evaluate overall cognition and come to similar conclusions as the ACE-3 and MoCA, and has the fidelity to identify dementing illnesses presenting in specific domains.^44^

### Accessibility of the ACoE

In creating the ACoE, we aimed to develop a software which is not only automated and remote, but also allows patients to answer naturalistically to mimic in-clinic testing. However, ACoE scores were negatively associated with age after removing the effect of cognition. We suspect this is due to the effect of decreasing technological skills with age.^58^ While we could not achieve perfect accessibility for the elderly we do develop an age-adjustment factor which removes the bias post-hoc. This adjustment is likely not perfect and will benefit from a larger sample size.

Aside from age, we found ethnicity, education, and employment did not influence scores. However, these demographic factors exert well-known biases against cognitive scores, especially the ACE-3 which is historically biased towards educated English speakers.^54,59–61^ It is possible we were not powered to find these effects. Further studies are needed to identify and control for other covariates.

### Application of the ACoE

We have specifically developed the ACoE to evaluate patients in a manner consistent with clinical practice. This aims to be a digital health tool which supports clinicians, not replace them. For this reason, we specifically developed the ACoE to return actual estimates of cognition and cognitive subdomains to aid interpretability and avoid utilizing ‘black-box’ algorithms.^62^ This can aid longitudinal tracking of patients, and while the ACoE does provide a patient classification based on these domains, clinicians have access to the raw results to exercise their own judgement.

Regarding the diagnostic classifications of the ACE-3, there are three potential classifications: confidently impaired (< 83%), indeterminate (84-88%), and confidently unimpaired (> 83%). Using these, the ACoE can help manage patient triaging.

Confidently impaired patients can be referred for treatment, while confidently unimpaired patients can be referred for annual testing and observation. Indeterminate patients (between 83%-89%) can be referred for in-person assessment for diagnostic clarification. This minimizes the load on clinician assessments of cognition.

It should be noted that the development of an ‘indeterminate’ region of the ACoE classification is not necessary statistically. Most similar cognitive examinations use a binary threshold, categorizing patients into “impaired” or “healthy”. However, this neglects the fact that the accuracy of their test varies across patient scores.^63^ Severely low scores strongly indicate impaired patients, while perfect scores strongly indicate intact patients. But what do we do with the in-between, where things are less clear? By directly acknowledging the existence of this uncertain region, we can build a tool which is less prone to mistakes^63–66^ and allows clinicians to critically evaluate outputs.^67^

### Limitations

The ACoE and this study are not without limitations. First, this study does not focus on individual etiologies causing dementia. Thus, it is possible that etiologies presenting in different cognitive domains may result in variable ACoE performance. However, our results do suggest that the ACoE is consistent in evaluation across cognitive domains compared to the ACE-3. This suggests it will evaluate as similarly as the ACE-3, and may simply fail where the ACE-3 fails.

The ACoE does require an internet connection. For patients with poor connections or poor hardware, ACoE performance may be hindered. If recordings of patients are poor, then the evaluations will be poor. We attempt to address this concern by primarily developing the ACoE with smartphones in mind, given the largely standardized hardware and focus on high quality microphones and touchscreens for daily use. Further smartphone also have reliable internet access, which helps address the other possible technical failure of low internet connectivity.

There are several other limitations of the ACoE. Language is an inherent bias of the ACoE. In attempting to create a naturalistic evaluation wherein a patient receives spoken instructions, answers with speech, and has language explicitly evaluated, we bias towards the language of development. Thus, in this case, the ACoE is limited to use in English speakers. Lastly, while our sample size was was guided by established requirements for the ICC, we understand the need for additional patients. However, in our current trial, additional sample would only increase precision of our significance estimates without directly assessing how the ACoE performs in different populations.

Further studies will need to evaluate the ACoE in specific patient populations such as Alzheimer Disease.

### Future Directions

There are many improvements to make to the ACoE. First, while this study was specifically guided by power analysis, a larger study is necessary. Having validated the remote validation, we aim to assess utility in specific etiologies and in larger sample sizes.

Second, the ACoE currently only evaluates *if* a question was answered correctly. Next, we will evaluate *how* patients answer, whether it is how they complete the clock drawing test or generate a list of words. This will enable development of further machine learning algorithms using the information within the answers themselves to aid in further diagnosis.

Lastly, the ACoE currently only focuses on if a foundational assessment of cognitively intact or cognitively impaired. While this is an important part of the initial evaluation of any patient, it is not enough. Future studies evaluating the ACoE in more neurodegenerative etiologies will allow development of specialized algorithms to aid in detection of specific diseases. For example, analysis of how a patient performs the clock drawing test will provide useful information relevant to posterior cortical atrophy, whereas data from speech will be critical for detecting the primary progressive aphasias.

## Data Availability

Data is available upon reasonable request.

## Acknowledgements/Conflicts/Funding Sources/Consent Statement

The authors would like to acknowledge Rachel Elizabeth Bethune Howard for support of the research.

Author CH is part of CogNet Inc., a company provided cognitive testing to rural Canadians.

There is no funding to disclose.

Subjects were recruited from the neurology and neuropsychology clinics, and all signed Institutional Review Board-approved consent forms. Substitute decision makers were included in the consent process of cognitively impaired patients. This work was conducted between 2022 and 2024.

## Notes

### Competing Interest Statement

The authors have declared no competing interest.

### Funding Statement

This study was funded by MITACS Accelerate

