## supplementary_figures for "The Autonomous Cognitive Examination: Machine-Learning Based Cognitive Examination"

$$ACoE\ Score = 80 + 19(Cognitive\ Status) - 0.22(Age - 30)$$

**Supplementary Equation 1**. The relationship of cognitive status on age upon ACoE scores, identified by multivariate regression.

$$Age\ Adjusted\ ACoE\ Score = ACoE\ Score + 0.22(Age - 30)$$

**Supplementary Equation 2.** The formula to adjust ACoE score for patient age, based on Supplementary Equation 1.

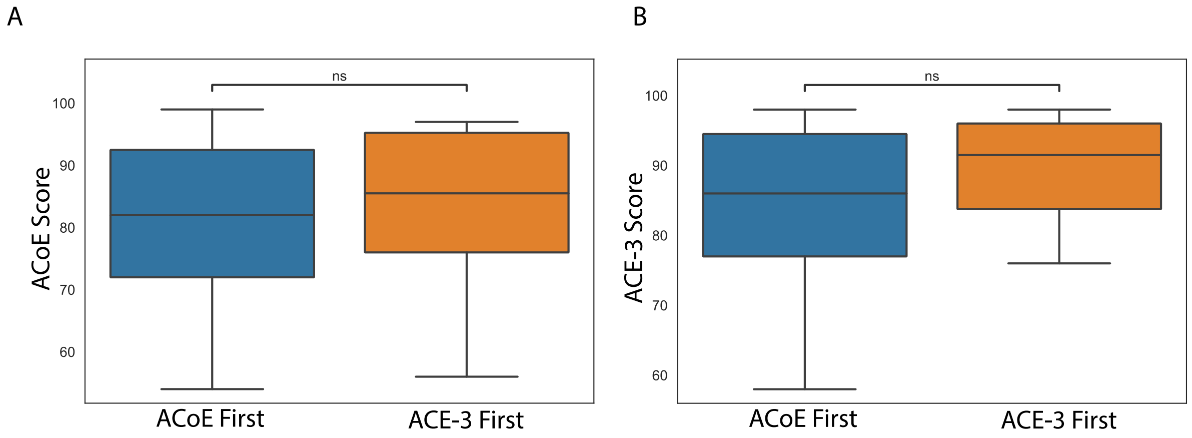


**Supplementary Figure 1.** A) Central tendencies of ACoE scores across randomization arms. Wilcoxon-Mann-Whitney U-Test demonstrates no significant difference (Group 1: 82, IQR 72.5-92; Group 2: 85.5, IQR 75-95.3, $p > 0.05$). B) Central tendencies of ACE-3 scores across randomization arms. Wilcoxon-Mann-Whitney U-Test demonstrates no significant difference (Group 1: 86, IQR (77-94.5); Group 2: 91.5, IQR 83.8-96, $p > 0.05$).

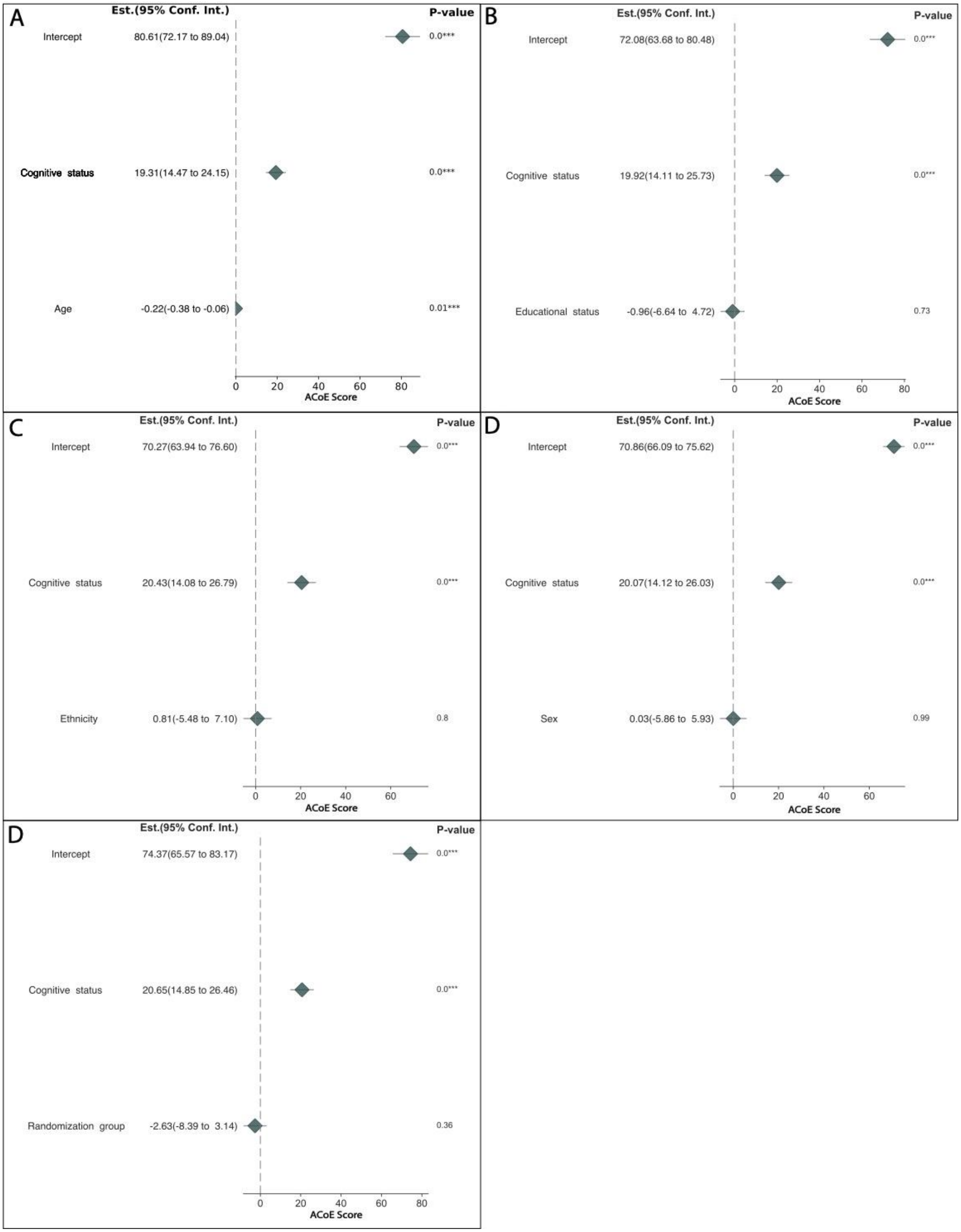


**Supplementary Figure 2**. **Only Age is Associated with ACoE Score After Controlling for Cognitive Status.** A) Multivariate regression of age and cognitive status upon ACoE score. B) Multivariate regression of educational status and cognitive status upon ACoE score. C)

Multivariate regression of ethnicity and cognitive status upon ACoE score. D) Multivariate regression of sex and cognitive status upon ACoE score. E) Multivariate regression of randomization group and cognitive status upon ACoE score. Each multivariate regression is presented as a forest plot, with coefficients reported as estimates with 95% confidence intervals. The coefficients and confidence intervals are forest plotted. P-values for each coefficient are shown at the right. Unstandardized coefficients are presented to ensure directly interpretability and application in relation to ACoE scores.

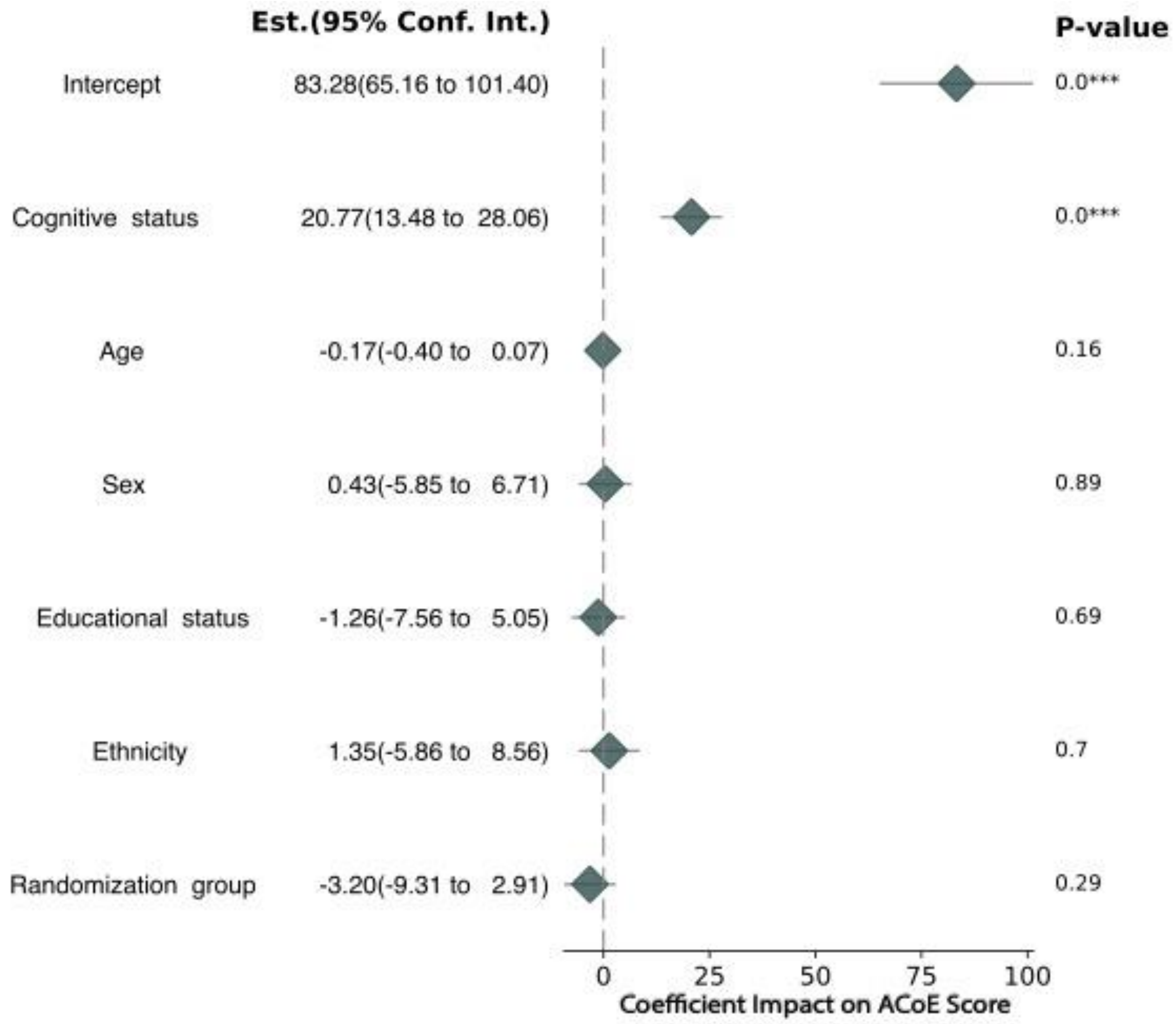


**Supplementary Figure 3. Only Cognitive Status is Associated with ACE-3 Score.** Multivariate regression of all covariates on ACE-3 scores. The coefficients and confidence intervals are forest plotted and presented as their estimates with 95% confidence intervals. P-values for each coefficient are shown at the right. Unstandardized coefficients are presented to ensure directly interpretability and application in relation to ACoE scores.

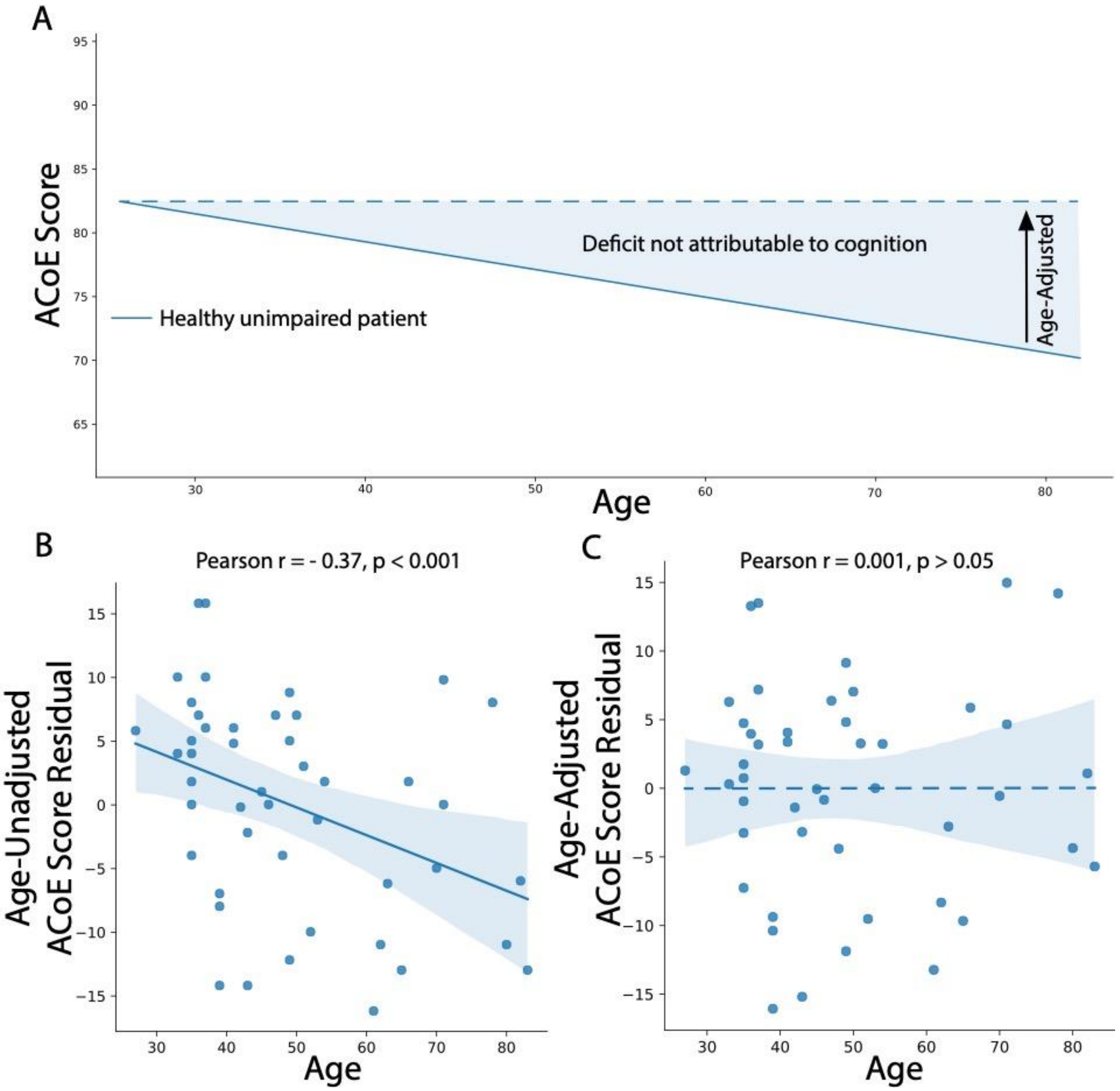


**Supplementary Figure 4. Adjustment of ACoE Scores for the Effect of Age.** A) A diagrammatic representation of the relationship between age and ACoE Scores. Even in cognitively intact patients, older age steadily reduced patient scores. B) Correlation of age to ACoE scores after regressing cognitive status out. There is a significant relationship between age and the residual ACoE scores, which is completely unrelated to cognition (Pearson $r = -0.37$, $p < 0.001$). C) Successful adjustment of ACoE scores for the effect of age. Correlation of age to the age-adjusted ACoE scores with cognitive status regressed out. After correcting for age, the relationship between age and residual ACoE scores (Pearson $r = 0.001$, $p > 0.05$).

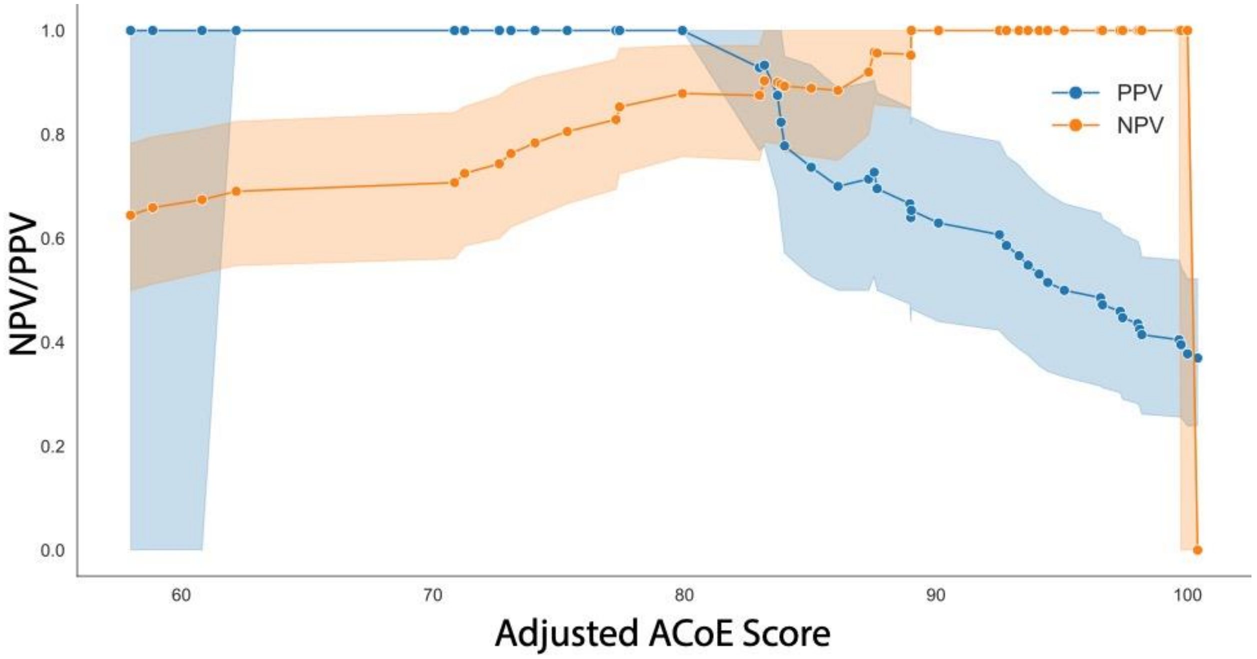


**Supplementary Figure 5. Score-specific positive and negative predictive values are consistent with the sensitivity and specificity of the ACoE.** Below a score of 83% corresponds to a positive predictive value of 0.93 (95CI 0.79-1.0). Above a score of 89% corresponds to a negative predictive value of 0.95 (95%CI 0.78-1.0). Negative and positive predictive values become unstable and meaningless at the extremes of data ranges, resulting in confidence intervals destabilizing and spanning 0.0-1.0.

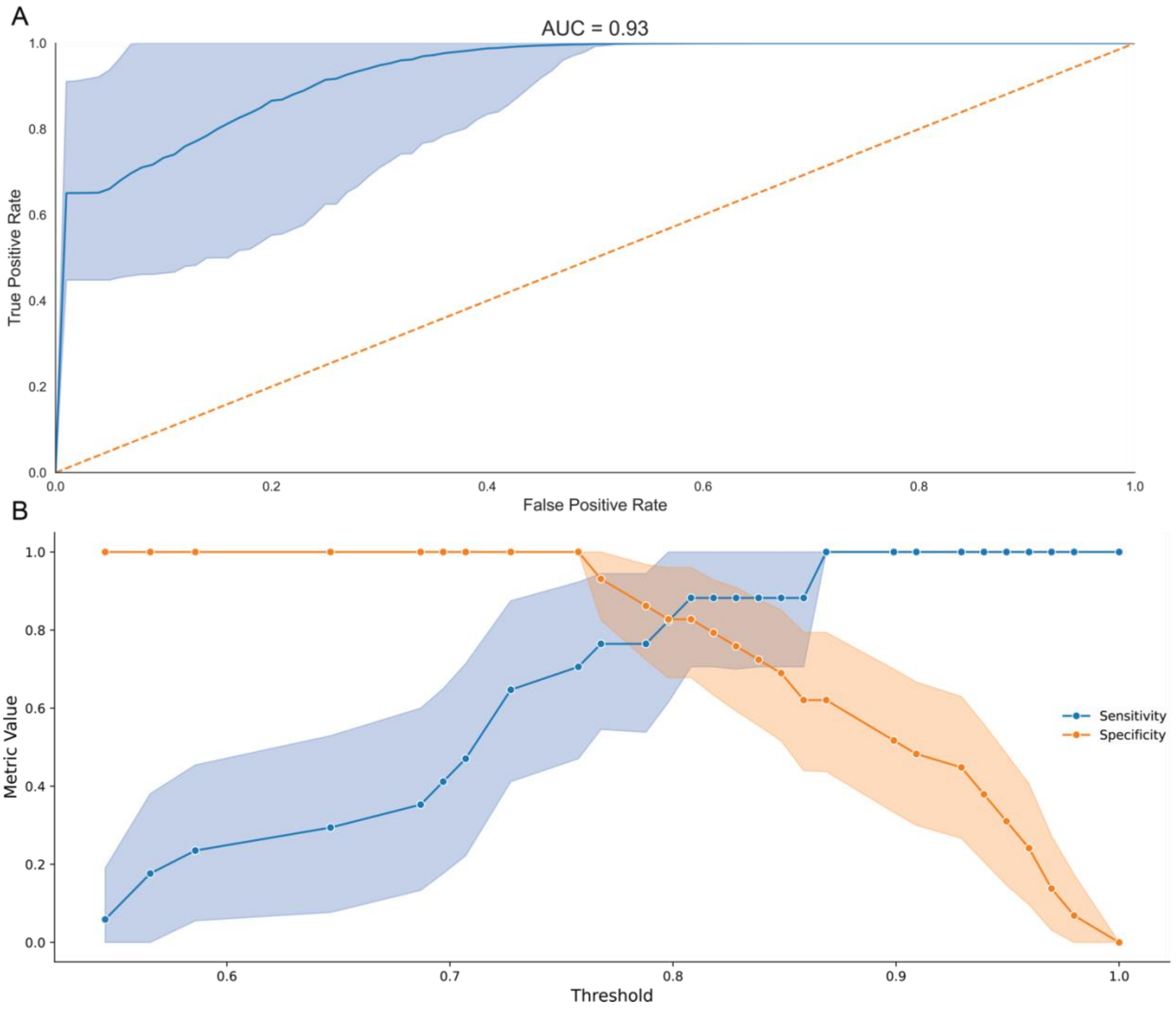


**Supplementary Figure 6**. **Unadjusted ACoE Score Maintains Diagnostic Accuracy of Paper-Based Tests.** A) ROC and AUC of the unadjusted ACoE. AUC of diagnosis is 0.93 compared to ACE-3 and MoCA evaluation of patients. B) Sensitivity and specificity curves across ACoE scores. Under an unadjusted score of 76%, the ACoE achieves a specificity of 1.0 (95%CI 1.0-1.0). Above an unadjusted score of 86%, the ACoE achieves a sensitivity of 1.0 (95%CI 1.0-1.0).

**Supplementary Table 1. Exam characteristics.** Each of the 19 primary questions is listed on the left, with the specific content of the question and the cognitive domain it is attributable to.

| Question Number | Question | Domain |
|---|---|---|
| 1 | Orientation to date, time, and method of testing. | Attention |
| 2 | Immediate recall of 3 words. | Attention |
| 3 | Serial subtraction of 7 from 100. | Attention |
| 4 | Delayed recall of 3 words. | Memory |
| 5 | Phonemic and semantic list generation. | Fluency |
| 6 | Immediate recall of 7 words. | Memory |
| 7 | Recall of semantic information. | Memory |
| 8 | Command-based comprehension. | Language |
| 9 | Written sentence. | Language |
| 10 | Single word repetition. | Language |
| 11 | Sentence repetition. | Language |
| 12 | Object naming. | Language |
| 13 | Object comprehension. | Language |
| 14 | Read words aloud. | Language |
| 15 | Object copy and clock drawing. | Visuospatial |
| 16 | Visual object counting. | Visuospatial |
| 17 | Obscured letter identification. | Visuospatial |
| 18 | Delayed recall of 7 words. | Memory |
| 19 | Recognition of 7 words. | Memory |

**Supplementary Table 2.** Cognitive Domain Central Tendencies Between ACoE and ACE-3. Values are reported as mean +/- standard error of the mean (median).

| Cognitive Domain | ACoE | ACE-3 | p-value |
|---|---|---|---|
| Attention | 16.11 +/- 0.32 (16.0) | 16.89 +/- 0.28 (18.0) | > 0.05 |
| Memory | 18.35 +/- 0.91 (19.0) | 20.6 +/- 0.77 (22.0) | > 0.05 |
| Fluency | 10.24 +/- 0.54 (11.0) | 11.23 +/- 0.43 (12.0) | > 0.05 |
| Language | 22.91 +/- 0.5 (24.0) | 23.77 +/- 0.5 (25.0) | > 0.05 |
| Visuospatial | 14.41 +/- 0.25 (14.0) | 14.63 +/- 0.36 (15.0) | > 0.05 |

**Supplementary Table 3.** Algorithm Scoring Central Tendencies Between ACoE and ACE-3. Values are reported as mean +/- standard error of the mean (median).

| Column Name | ACoE | ACE-3 | p-value |
|---|---|---|---|
| Convolutional Neural Network | 6.89 +/- 0.16 (6.5) | 7.03 +/- 0.26 (8.0) | > 0.05 |
| Natural Language Processing | 46.89 +/- 1.27 (47.0) | 50.26 +/- 1.09 (52.0) | > 0.05 |
| Expert Algorithm | 28.24 +/- 0.64 (30.0) | 29.83 +/- 0.48 (31.0) | > 0.05 |

**Supplementary Table 4.** Question Scoring Central Tendencies Between ACoE and ACE-3. Values are reported as mean +/- standard error of the mean (median).

| Question | ACoE | ACE-3 | p-value |
|---|---|---|---|
| 1 | 9.63 +/- 0.11 (10.0) | 9.69 +/- 0.15 (10.0) | > 0.05 |
| 2 | 2.54 +/- 0.13 (3.0) | 2.97 +/- 0.03 (3.0) | > 0.05 |
| 3 | 3.93 +/- 0.24 (5.0) | 4.23 +/- 0.21 (5.0) | > 0.05 |
| 4 | 2.35 +/- 0.15 (3.0) | 2.71 +/- 0.11 (3.0) | > 0.05 |
| 5 | 10.24 +/- 0.54 (11.0) | 11.23 +/- 0.43 (12.0) | > 0.05 |
| 6 | 5.09 +/- 0.33 (6.0) | 5.97 +/- 0.22 (7.0) | > 0.05 |
| 7 | 2.72 +/- 0.15 (3.0) | 3.03 +/- 0.15 (3.0) | > 0.05 |
| 8 | 2.43 +/- 0.11 (3.0) | 2.89 +/- 0.05 (3.0) | > 0.05 |
| 9 | 1.59 +/- 0.1 (2.0) | 1.86 +/- 0.06 (2.0) | > 0.05 |
| 10 | 1.78 +/- 0.07 (2.0) | 1.8 +/- 0.07 (2.0) | > 0.05 |
| 11 | 1.83 +/- 0.06 (2.0) | 1.89 +/- 0.05 (2.0) | > 0.05 |
| 12 | 11.0 +/- 0.32 (12.0) | 11.14 +/- 0.3 (12.0) | > 0.05 |
| 13 | 3.74 +/- 0.09 (4.0) | 3.6 +/- 0.12 (4.0) | > 0.05 |
| 14 | 0.54 +/- 0.07 (1.0) | 0.6 +/- 0.08 (1.0) | > 0.05 |
| 15 | 6.89 +/- 0.16 (6.5) | 7.03 +/- 0.26 (8.0) | > 0.05 |
| 16 | 3.76 +/- 0.09 (4.0) | 3.8 +/- 0.1 (4.0) | > 0.05 |
| 17 | 3.76 +/- 0.1 (4.0) | 3.8 +/- 0.12 (4.0) | > 0.05 |
| 18 | 4.0 +/- 0.36 (4.0) | 4.34 +/- 0.44 (5.0) | > 0.05 |
| 19 | 4.2 +/- 0.18 (5.0) | 4.54 +/- 0.13 (5.0) | > 0.05 |
